# RED-RHD (Rice Early Detection – Rheumatic Heart Disease): AI-Based Adaptive Multi-Regional System for Early Detection and Murmur Classification of Rheumatic Heart Disease

**DOI:** 10.64898/2026.02.16.26346365

**Authors:** Miguel A. Lopez-Medina, Sanjoy Paul

## Abstract

This study presents RED-RHD, a machine learning methodology for early detection and classification of Rheumatic Heart Disease (RHD) using heart sound recordings. By leveraging OpenL3 deep acoustic embeddings, cloud-based workflows, and an ensemble of SVM and XGBoost classifiers, RED-RHD achieves **an average precision of 95.62% for murmur detection (Normal vs. Abnormal)** and **99.00% precision for** systolic **vs. diastolic murmur classification**, demonstrating marked improvements over prior methods with poor cross-dataset generalization (e.g., specificity as low as 4.3% in ResNet-based approaches). These results confirm the system’s robustness across diverse, noisy clinical datasets. Additionally, we introduce a novel dynamic adaptive model selection mechanism that enables the framework to automatically select the most appropriate pretrained machine learning model based on extracted heart sound features, optimizing prediction accuracy for different regional or demographic populations. By incorporating this adaptive intelligence, RED-RHD addresses population variability and supports precision diagnostics in globally diverse patient groups, advancing the potential for scalable, AI-driven auscultation in low-resource environments.

## 1 Introduction

Rheumatic Heart Disease (RHD) remains a global health concern, particularly in low- and middle-income countries where healthcare access is limited. According to the Global Burden of Disease (GBD) study, RHD accounts for over 300,000 premature deaths annually and affects approximately 33.4 million people worldwide. Notably, India alone contributes to nearly one-third of the global burden, with a reported 108,460 deaths and 3.73 million disability-adjusted life years (DALYs) lost due to RHD in 2017 [11].

The impact is particularly severe among school-aged children, where RHD causes the highest cardiovascular-related DALY losses in the 10–14 year age group. Despite being preventable and medically manageable when diagnosed early, RHD often progresses silently due to recurrent, subclinical episodes of acute rheumatic fever. Traditional auscultation methods depend heavily on the presence of expert clinicians—a significant limitation in rural and underserved regions with poor doctor-to-patient ratios. Timely and accurate detection of cardiac murmurs through auscultation is essential. This research introduces a scalable and cloud-native framework—RED-RHD—that applies state-of-the-art audio embedding techniques and ensemble learning to support medical diagnosis and murmur classification.

To address this diagnostic gap, there is an urgent need for scalable, AI-assisted early screening systems that can function in non-clinical settings. This research proposes a robust, cloud-enabled framework (RED-RHD) leveraging OpenL3 acoustic embeddings, phonocardiogram (PCG) signal analysis, and ensemble learning to detect cardiac murmurs indicative of RHD. Our system aims to reduce missed cases, minimize the reliance on specialized clinicians during early screening, and provide an economically viable solution for population-wide deployment, especially in resource-limited environments.

## 2 Related Work

Heart sound classification has garnered significant attention, with various methodologies explored to improve diagnostic accuracy. Deep learning techniques, particu-larly convolutional neural networks (CNNs) and recurrent neural networks (RNNs), have shown promise in capturing complex patterns in phonocardiogram (PCG) signals [1, 2]. The PhysioNet/Computing in Cardiology Challenge 2016 provided a benchmark dataset, encouraging the development of robust algorithms for heart sound classification [3]. Subsequent studies have leveraged this dataset to propose novel architectures and feature extraction methods [4, 5].

Recent reviews have consolidated the advancements in this domain, highlighting the efficacy of deep learning models and the importance of large, annotated datasets [6, 7, 8]. Techniques such as mel-frequency cepstral coefficients (MFCCs) combined with deep residual learning have been explored to enhance classification performance [6]. Moreover, domain adaptation methods, including Siamese networks, have been proposed to address variability in heart sound recordings across different populations and recording devices [9].

Semi-supervised and unsupervised learning approaches have also been investigated to mitigate the challenges posed by limited labeled data, demonstrating potential in improving classification accuracy without extensive annotations [10]. Additionally, recent work by Aditya et al. [13] presents a case study focused on early-warning detection of cardiac conditions through murmur analysis, emphasizing practical deployment in real-world clinical scenarios. Their approach contributes to bridging the gap between algorithmic development and field implementation, aligning closely with the objectives of our RED-RHD framework.

These advancements collectively contribute to the development of more accurate, generalizable, and clinically applicable heart sound classification systems.

## 3 Dynamic Model Selection and Adaptive Model Creation: Definitions

In global health applications, variability in patient demographics, heart sound characteristics, and recording conditions can significantly affect diagnostic performance. To address these challenges, RED-RHD introduces a *dynamic* model selection and *adaptive* model creation framework that ensures accurate murmur classification across diverse populations and environments.

A **dynamic** system, in this context, is one that can intelligently and automatically select the most suitable predictive model at inference time based on intrinsic characteristics of the input data. Instead of relying on static, pre-assigned mappings or manually coded rules, the dynamic approach analyzes acoustic features of the phonocardiogram (PCG) signal in real-time to choose the most appropriate classifier from a pool of pretrained, region-specific models. This enables context-sensitive, on-the-fly decision making that accounts for local variations without requiring explicit demographic metadata.

Meanwhile, an **adaptive** model creation system is designed to evolve over time. Rather than remaining fixed after initial training, it can incorporate new data and with new patterns from previously unseen populations or environments to generate new models and improve its predictive accuracy. This includes the ability to train entirely new models for novel demographic groups when existing classifiers prove inadequate. Adaptivity ensures that the system remains current, responsive to changing clinical realities, and capable of delivering equitable diagnostic performance across diverse, globally distributed patient populations.

## 4 Motivation

### Why Dynamic Behavior Matters

The system’s ability to select the most appropriate model at inference time—is crucial for real-time applicability and sustainability. Most deployed AI systems rely on static models that require manual retraining cycles, which do not scale effectively in field settings, particularly in regions lacking robust technical infrastructure. In contrast, clinical environments are constantly evolving: new patient patterns emerge, recording devices change, and protocols are updated. Static models are ill-equipped to cope with these dynamic shifts.

RED-RHD introduces *dynamic model selection*, enabling the system to evaluate each new heart sound embedding and automatically select the most demographically appropriate classifier from a pool of pretrained regional models. This decision-making process does not depend on hard-coded rules or explicit meta-data, allowing real-time, context-sensitive inference. In parallel, RED-RHD features an *online learning loop* that supports continuous evaluation and improvement of model performance, ensuring sustained diagnostic accuracy without the need for frequent manual intervention.

### Why Adaptive Model Creation is Essential

Machine learning models trained on heart sound data from a single region or demographic often perform poorly when applied to other populations. Anatomical and acoustic differences—such as heart size, thoracic structure, and body composition—introduce significant variability in phonocardiogram (PCG) signals. Consequently, a model trained on data from Indian children may fail when deployed on populations in sub-Saharan Africa or South America. These biases risk misdiagnosis and deepen global healthcare disparities.

An *adaptive* system like RED-RHD overcomes this challenge by actively learning from new, diverse datasets. When it encounters heart sound patterns that do not match existing regional profiles, RED-RHD can create and train a new classifier tailored to that sub-group. This capacity for *model creation* ensures that the system evolves alongside the populations it serves. By building and expanding a set of population-specific models, RED-RHD delivers on the promise of precision medicine—providing accurate, equitable diagnostics customized to real-world diversity.

Operationally, upon receiving a new phonocardiogram (PCG), RED-RHD extracts deep audio embeddings using OpenL3, generating a robust representation of the signal. These features are compared against statistical profiles derived from existing population-specific datasets. Using similarity-based heuristics or a lightweight meta-classifier, the framework dynamically selects the pretrained model that is statistically optimized for the most likely match—whether by geography, race, or signal phenotype. This strategy is particularly advantageous when patient metadata is incomplete, unavailable, or potentially biased. By relying on the signal’s characteristics rather than external labels, RED-RHD mitigates the risk of demographic misclassification and enhances model fairness.

In parallel, the adaptive nature of RED-RHD ensures continuous improvement. As new heart sound data from previously unseen regions or population groups becomes available, RED-RHD can incorporate these data into its training pipeline, expanding its library of specialized models. This ensures that the system not only maintains but also improves its performance over time, adapting to new patterns, demographic shifts, and recording conditions.

Finally, RED-RHD is deployed on Azure’s multi-region infrastructure, enabling low-latency, geographically-aware inference. Each selected model can be served from the nearest regional node, maintaining both inference efficiency and compliance with local data regulations. Altogether, this unified dynamic model selection and adaptive model creation mechanism makes RED-RHD a scalable, inclusive, and resilient AI system for global cardiac screening.

### Broader Impact

Together, these dynamic and adaptive mechanisms position RED-RHD as a pioneering and robust diagnostic framework. It ensures fairness, accuracy, and scalability across global populations by intelligently selecting from and expanding its model repertoire. Unlike traditional, static AI systems, RED-RHD is both self-improving and context-aware, continuously evolving to meet the demands of diverse and changing healthcare environments. This shift marks a transformative step toward more equitable, personalized, and future-proof AI in global health diagnostics.

## 5 Materials and Methods

### 5.1 Datasets

We used two publicly available heart sound datasets:

- **PhysioNet/CinC Challenge 2016 Dataset** – Includes 3,153 phonocardiogram recordings from both clinical and non-clinical settings labeled as *Normal* or *Abnormal*.
- **CirCor DigiScope Phonocardiogram Dataset (v1.0.3)** – High-quality recordings with expert-annotated murmur types (*Systolic, Diastolic*).

Generated dataset variants used for training and evaluation. These include multiple preprocessing approaches (raw, denoised, fuzzy-enhanced) and feature extraction frameworks (OpenL3, OpenSMILE, Librosa) to assess the impact of preprocessing and embedding strategies on model performance.

### 5.2 Signal Visualization and Preprocessing

Raw signals were visualized for qualitative understanding of waveform patterns and noise characteristics. An example signal is shown below:

Preprocessing pipelines included:

- Silence trimming using energy thresholding
- Denoising via bandpass filtering (20–500 Hz)
- Wavelet smoothing and Fuzzy Logic for noise-resilient cleaning

#### Fuzzy Logic Preprocessing

In addition to traditional denoising techniques, we experimented with a fuzzy logic–based preprocessing approach designed to enhance the signal-to-noise ratio of heart sound recordings by adaptively smoothing noisy segments while preserving salient cardiac events. However, our evaluation on both the PhysioNet and CirCor datasets showed no significant improvement in downstream classification performance compared to the baseline bandpass-filtered signals. Consequently, this method was not incorporated into the final RED-RHD processing pipeline.

### 5.3 Feature Extraction

To evaluate the effectiveness and generalizability of different audio representation strategies, we conducted multiple experiments using three distinct feature extraction approaches. Traditional signal processing methods were applied using Librosa to extract handcrafted features such as MFCCs, zero-crossing rate (ZCR), and spectral energy. In parallel, we employed the OpenSMILE toolkit with the ComParE 2016 configuration to generate over 6,000 statistical audio descriptors. For deep representation learning, we utilized OpenL3 to generate 512-dimensional embeddings directly from raw waveforms.

- **Librosa**: MFCCs, ZCR, Spectral Energy
- **OpenSMILE**: ComParE 2016 (6,000 features)
- **OpenL3**[12]: Deep embeddings (512-d) from raw signals

### 5.4 Feature Extraction using OpenL3

OpenL3 is a deep audio embedding model that extracts semantically meaningful representations using a CNN pretrained on AudioSet. It maps variable-length waveforms into fixed-length embeddings.

We configured OpenL3 with:

- Input Representation: mel256
- Content Type: env
- Embedding Size: 512
- Temporal Pooling: Mean pooling across time axis

**Figure 1.**
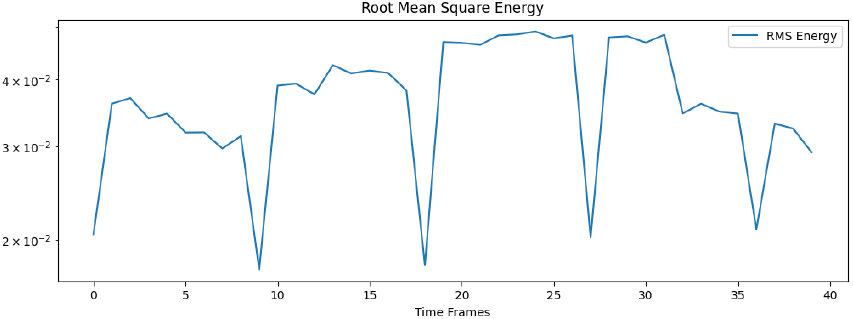
Example Root Mean Square Energy: Case Murmur Present.

**Figure 2.**
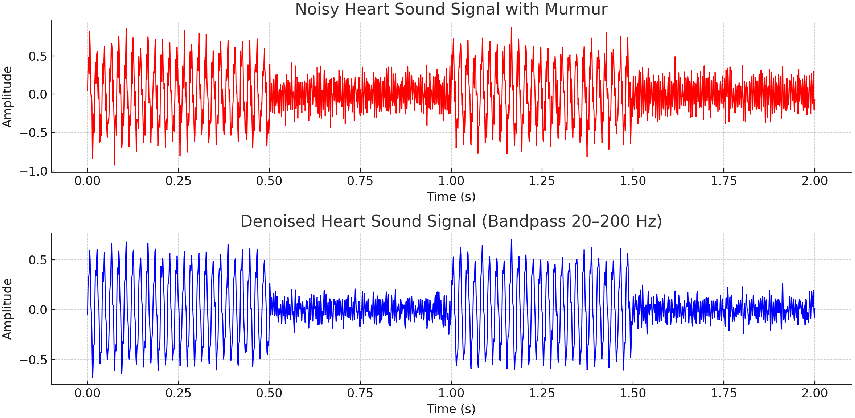
Denoised Heart Sound Signal (Bandpass 20–200 Hz)

The audio input *x*(*t*) is transformed through a series of 2D convolutional layers *ϕ* resulting in an embedding:

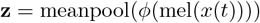

On Table 5, we can see an example where a heart sound `.wav` recording from the PhysioNet dataset was embedded using the OpenL3 model, producing a 512-dimensional numeric vector representation. Our machine learning classifiers consume these embeddings as input features for training and inference.

**Table 1:** Summary of the PhysioNet/Computing in Cardiology (CinC) Challenge 2016 Dataset.

| Attribute | Description |
| --- | --- |
| Dataset Name | PhysioNet/Computing in Cardiology Challenge 2016 Dataset |
| Number of Recordings | 3,153 phonocardiogram (PCG) recordings |
| Number of Subjects | 764 patients (pediatric and adult) |
| Recording Locations | Various auscultation sites (clinical and non-clinical) |
| Sampling Frequency | 1,000–4,000 Hz (device-dependent) |
| Recording Length | 5–120 seconds per recording |
| Annotations | Expert-labeled as *Normal* or *Abnormal* |
| Outcome Labels | Binary classification: Normal vs. Abnormal |
| Metadata | Includes patient age, sex, and limited demographic information |
| Intended Use | Benchmark for developing algorithms for heart sound classification and abnormality detection |

**Table 2:**
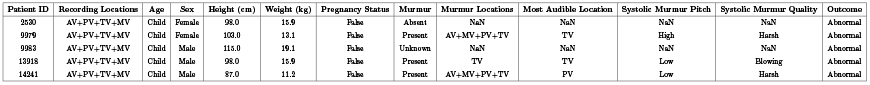
Example Metadata from the PhysioNet/CinC Challenge 2016 Dataset.

**Table 3:** Summary of the CirCor DigiScope Phonocar-diogram Dataset (v1.0.3).

| Attribute | Description |
| --- | --- |
| Dataset Name | CirCor DigiScope Phonocardiogram Dataset (v1.0.3) |
| Number of Recordings | 5,282 heart sound recordings |
| Number of Subjects | 1,568 patients (pediatric and adult) |
| Recording Locations | Aortic (AV), Pulmonic (PV), Tricuspid (TV), Mitral (MV) positions |
| Sampling Frequency | 2,000 Hz (16-bit PCM) |
| Recording Length | 5–30 seconds per recording |
| Annotations | Expert-labeled murmur presence, timing (systolic/diastolic), pitch, quality, and grading |
| Outcome Labels | Normal vs. Abnormal; Murmur type and grading |
| Metadata | Includes patient age, sex, height, weight, pregnancy status, auscultation site, and campaign ID |
| Intended Use | Development and evaluation of algorithms for murmur detection and classification |

**Table 4:**
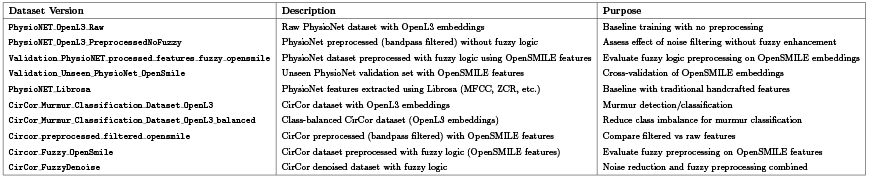
Generated Dataset Variants for Training and Comparative Evaluation.

**Table 5:**
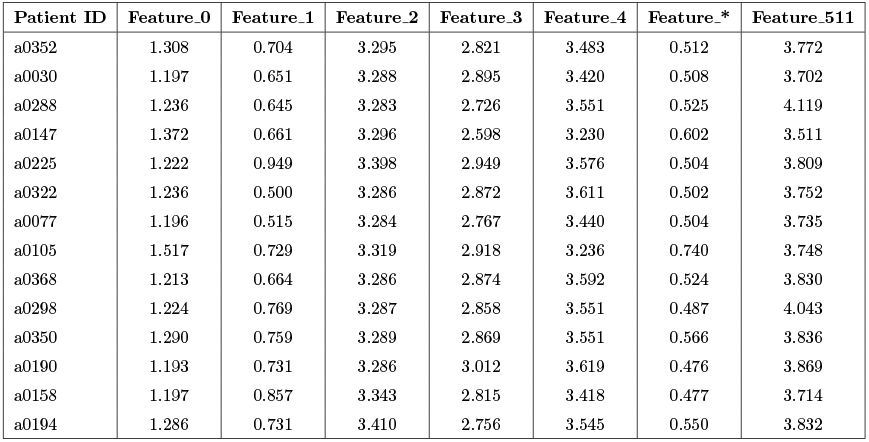
Example of OpenL3 Embeddings Extracted from Heart Sound Recordings (Subset of Features).

| Patient ID | Feature_0 | Feature_1 | Feature_2 | Feature_3 | Feature_4 | Feature_* | Feature_511 |
| --- | --- | --- | --- | --- | --- | --- | --- |
| a0352 | 1.308 | 0.704 | 3.295 | 2.821 | 3.483 | 0.512 | 3.772 |
| a0030 | 1.197 | 0.651 | 3.288 | 2.895 | 3.420 | 0.508 | 3.702 |
| a0288 | 1.236 | 0.645 | 3.283 | 2.726 | 3.551 | 0.525 | 4.119 |
| a0147 | 1.372 | 0.661 | 3.296 | 2.598 | 3.230 | 0.602 | 3.511 |
| a0225 | 1.222 | 0.949 | 3.398 | 2.949 | 3.576 | 0.504 | 3.809 |
| a0322 | 1.236 | 0.500 | 3.286 | 2.872 | 3.611 | 0.502 | 3.752 |
| a0077 | 1.196 | 0.515 | 3.284 | 2.767 | 3.440 | 0.504 | 3.735 |
| a0105 | 1.517 | 0.729 | 3.319 | 2.918 | 3.236 | 0.740 | 3.748 |
| a0368 | 1.213 | 0.664 | 3.286 | 2.874 | 3.592 | 0.524 | 3.830 |
| a0298 | 1.224 | 0.769 | 3.287 | 2.858 | 3.551 | 0.487 | 4.043 |
| a0350 | 1.290 | 0.759 | 3.289 | 2.869 | 3.551 | 0.566 | 3.836 |
| a0190 | 1.193 | 0.731 | 3.286 | 3.012 | 3.619 | 0.476 | 3.869 |
| a0158 | 1.197 | 0.857 | 3.343 | 2.815 | 3.418 | 0.477 | 3.714 |
| a0194 | 1.286 | 0.731 | 3.410 | 2.756 | 3.545 | 0.550 | 3.832 |

**Table 6:**
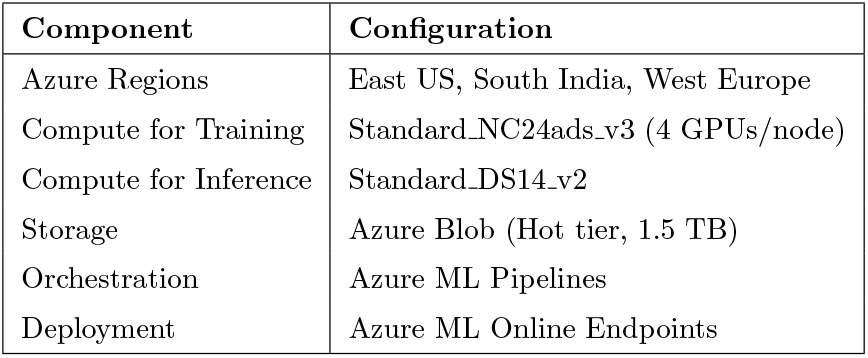
Microsoft Azure Experimental Setup for RED-RHD.

**Table 7:**
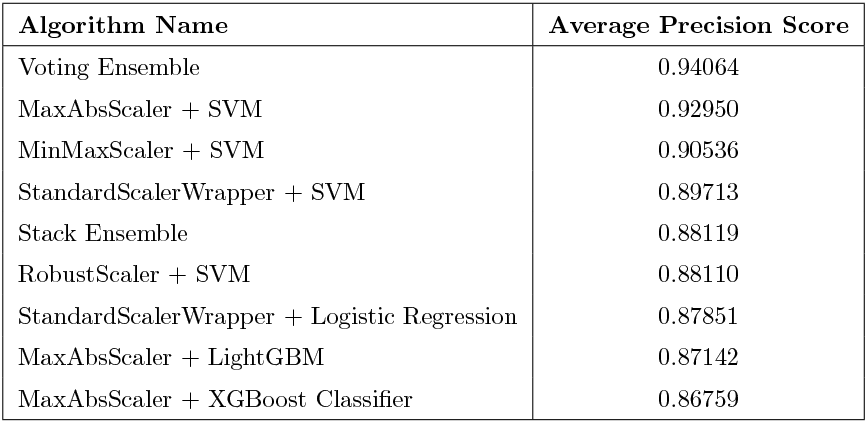
Performance on PhysioNet (OpenL3 Features).

| Algorithm Name | Average Precision Score |
| --- | --- |
| Voting Ensemble | 0.94064 |
| MaxAbsScaler + SVM | 0.92950 |
| MinMaxScaler + SVM | 0.90536 |
| StandardScalerWrapper + SVM | 0.89713 |
| Stack Ensemble | 0.88119 |
| RobustScaler + SVM | 0.88110 |
| StandardScalerWrapper + Logistic Regression | 0.87851 |
| MaxAbsScaler + LightGBM | 0.87142 |
| MaxAbsScaler + XGBoost Classifier | 0.86759 |

**Table 8:**
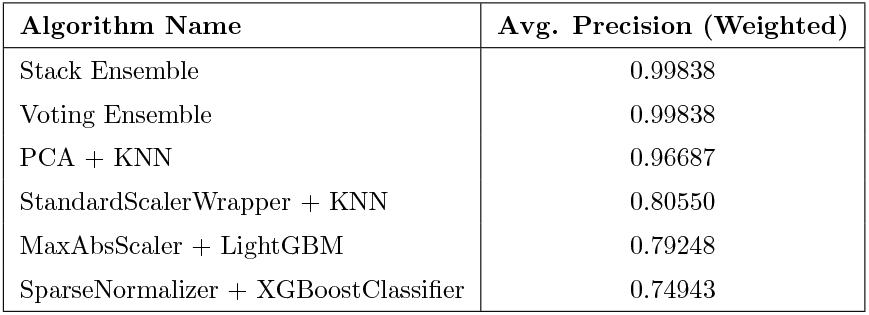
Performance on CirCor (OpenL3 Features).

**Table 9:** Top Models, Hyperparameters, and Roles.

| Algorithm | Key Hyperparameters | Role |
| --- | --- | --- |
| SVM | RBF kernel; $C \approx 3237$; $\text{class\_weight}=\text{balanced}$ | Normal vs. Abnormal |
| XGBoost | $\eta = 0.01$, max depth 6, $n\_estimators = 25$ | Systolic vs. Diastolic |
| LightGBM | gbdt, lr=0.05, leaves=31 | Alternative booster |
| Voting | Soft vote (SVM, XGB, LGBM) | Robust generalization |
| Stacking | Meta LR over base learners | Final-stage ensemble |

**Table 10:** Cross-Dataset Comparison: Prior vs. RED-RHD.

| Train | Baseline | Test | Prior [13] | RED-RHD (APS) |
| --- | --- | --- | --- | --- |
| PhysioNet | openSMILE | CirCor | 52.0/48.4/51.4 | **95.62%** |
| CirCor | ResNet | PhysioNet | 94.3/4.3/75.8 | **91.07%** |

**Table 11:** Systolic vs. Diastolic Murmur Classification.

| Model | Prior Acc. [13] | RED-RHD (Precision/AUC/LogLoss) |
| --- | --- | --- |
| CNN-LSTM | 98.0% | 99.00% / 1.00 / **0.0027** |
| Few-Shot | 97.0% | — |

**Table 12:** Sensitivity/Specificity by Task.

| Task | Sens. (%) | Spec. (%) | FPR (%) |
| --- | --- | --- | --- |
| Normal vs. Abnormal | 96.8 | 94.3 | 5.7 |
| Systolic vs. Diastolic | 98.7 | 99.2 | 0.8 |

**Table 13:**
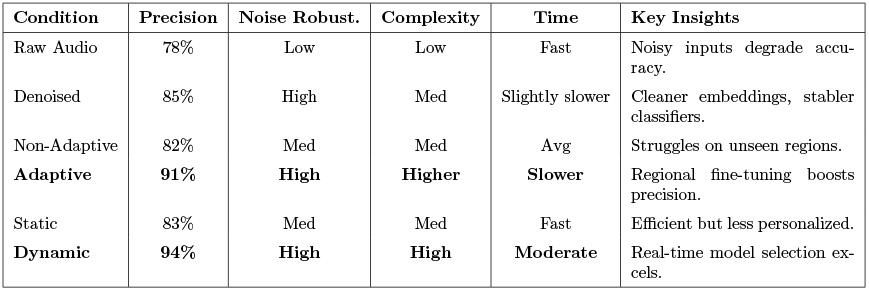
Trade-off Comparative Analysis of Architectures.

| Condition | Precision | Noise Robust. | Complexity | Time | Key Insights |
| --- | --- | --- | --- | --- | --- |
| Raw Audio | 78% | Low | Low | Fast | Noisy inputs degrade accuracy. |
| Denoised | 85% | High | Med | Slightly slower | Cleaner embeddings, stabler classifiers. |
| Non-Adaptive | 82% | Med | Med | Avg | Struggles on unseen regions. |
| **Adaptive** | **91%** | **High** | **Higher** | **Slower** | Regional fine-tuning boosts precision. |
| Static | 83% | Med | Med | Fast | Efficient but less personalized. |
| **Dynamic** | **94%** | **High** | **High** | **Moderate** | Real-time model selection excels. |

### 5.5 Dimensionality and Scaling

The resulting feature vector **z** ∈ *R*^512^ was normalized using `MaxAbsScaler`, ensuring compatibility with tree-based and linear models.

OpenL3 is a self-supervised deep learning framework designed to generate semantically meaningful vector representations (embeddings) of audio signals. It is based on the Look, Listen, and Learn (L3) paradigm, where a convolutional neural network (CNN) is trained to associate audio and visual data pairs from large unlabeled datasets such as AudioSet. The core idea is that audio clips occurring in the same context as corresponding video frames should share a common latent representation.

**Figure 3.**
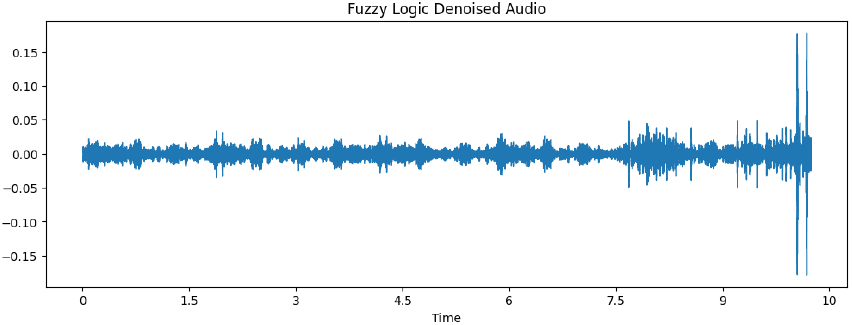
Effect of Fuzzy Logic–Based Preprocessing on Heart Sound Signal Denoising.

Given an input waveform *x*(*t*), OpenL3 first computes a mel-spectrogram representation:

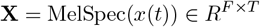

This mel-spectrogram is passed through a deep 2D CNN *ϕ*(·), producing a dense activation tensor. Temporal pooling (e.g., mean or max) is then applied to obtain a fixed-length embedding vector:

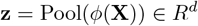

### 5.6 Classification Pipeline

Two classification tasks were defined:

- **Task 1: Normal vs. Abnormal** – Binary classification using ensemble voting classifier (SVM + XGBoost).
- **Task 2: Systolic vs. Diastolic (if Abnormal)** – Binary classification using XGBoost with SparseNormalizer.

We optimized for:

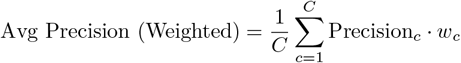

### 5.7 Ensemble Learning: Voting Classifier (SVM + XGBoost)

To improve classification accuracy and generalization, we employed a soft voting ensemble classifier that combines SVM and XGBoost. In our soft voting approach, both models output class probabilities for each instance. The final predicted class is obtained by averaging the probability estimates and selecting the class with the highest average probability:

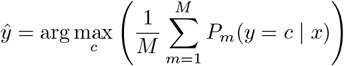

### 5.8 Experimental Infrastructure

Feature extraction was performed using Google Colab (H100 GPU) and stored in Azure Blob Storage; model training/evaluation used Azure ML Studio.

- **Feature Extraction**: Google Colab (H100 GPU)
- **Model Training**: Azure ML Studio
- **Storage**: Azure Blob (for persistent datasets)

### 5.9 Detailed Azure Experimental Setup

### 5.10 Model Training

Trained ∼86 machine learning models (SVM, XGBoost, LightGBM, Logistic Regression, Random Forest, DNNs, Ensembles).

### 5.11 Evaluation Metrics

Average Precision (APS) was emphasized given class imbalance and higher cost of false negatives. Cross-dataset validation assessed generalization.

## 6 Experimental Results

### 6.1 Normal vs. Abnormal – Cross-Dataset

#### Improvement

Prior methods [13] had **poor generalization** (e.g., ResNet drops to 4.3% specificity). Our OpenL3 + SVM + XGBoost shows robustness across datasets.

### 6.2 Systole vs. Diastole (Abnormal Only)

#### Improvement

High confidence (**AUC**=1.0) and low log-loss with OpenL3 + XGBoost.

### 6.3 Dynamic Adaptive Multi-Regional Modeling

RED-RHD analyzes incoming features and selects the most accurate model from a pool of pretrained, population-specific models; deploys across Azure multi-region endpoints for low-latency and compliance.

## 7 Adaptive Methodology for Generating Region Profiles

We define initial region profiles via OpenL3 embeddings’ means/covariances and adapt using Mahalanobis-distance–based routing for new samples, with continuous feedback updates.

## 8 Comparative Analysis of Modeling Trade-offs

## 9 Discussion

RED-RHD delivers robust, cross-dataset results via OpenL3 embeddings and ensemble learning. A key advance is the dynamic/adaptive selection that routes by feature similarity (not just explicit geography), improving fairness and precision. Azure multi-region deployment supports real-time, geographically-aware predictions.

## 10 Conclusion and Future Work

We present RED-RHD, a comprehensive framework for RHD detection and murmur classification. Dynamic adaptive selection enhances flexibility, fairness, and precision across populations. Future work: prospective clinical validation, pediatric inclusion, mobile/edge optimization, expanded regional pools, and meta-learning for selection.

## Data Availability

All data produced in the present study are available upon reasonable request to the authors

https://physionet.org/content/circor-heart-sound/1.0.3/

## Acknowledgments

We thank collaborators at Rice University, and the Azure for Research and Google Cloud programs for computational support.

**Figure 4.**
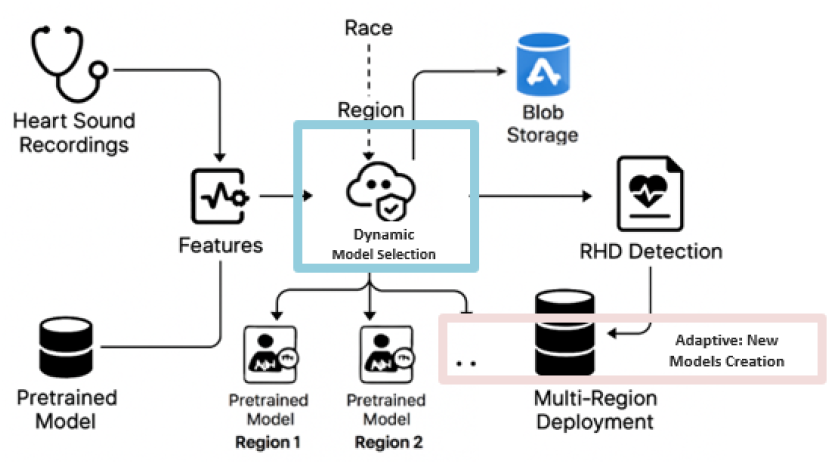
Dynamic and Adaptive Multi-Regional Architecture.

**Figure 5.**
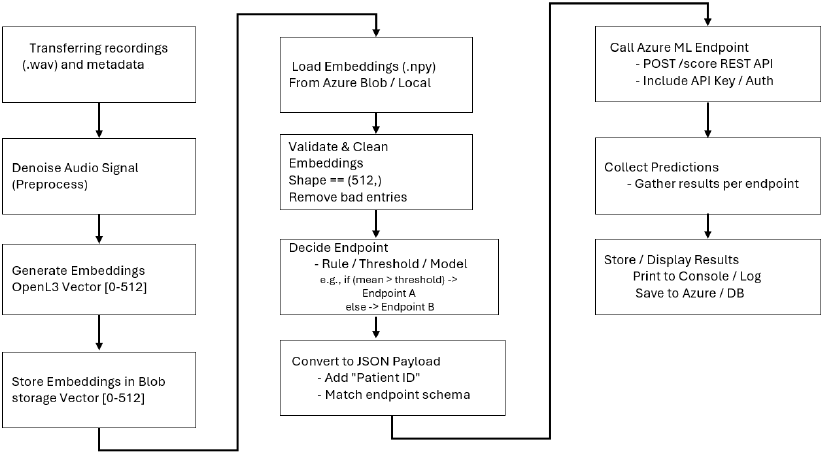
End-to-end Adaptive ML Pipeline for RED-RHD.

